# Polysubstance use: Delay discounting in relationship to remission

**DOI:** 10.1101/2025.10.01.25337100

**Authors:** Fatima Quddos, Devin C. Tomlinson, Rafaela M. Fontes, Allison N. Tegge, Warren K. Bickel

## Abstract

**Background:** Remission from substance use disorders (SUDs) is typically conceptualized as an all-or-none phenomenon. Here, we present a novel construct: proportion of remission (PrR; i.e., the proportion of substances an individual is in remission from relative to lifetime SUD history), a continuous construct that may better capture progress towards recovery in polysubstance use.

**Methods:** Individuals (n= 2,417) in recovery from SUDs were recruited from International Quit and Recovery Registry (IQRR). Individuals completed a $1000 adjusting amount delay discounting (DD) task, and questions about current and past substance use over the past 12 months and lifetime. We estimated a series of single-level binomial regressions models using PrR as the independent variable and DD, maximum time in recovery, and maximum quit time as dependent variables. In addition, we performed a moderated mediation analysis to understand the relationship between PrR and recovery variables.

**Results:** We report that DD, maximum time in recovery, and maximum quit time significantly predicted PrR in individuals with a history of polysubstance use. Further, we found that the relationship between maximum time in recovery and PrR was mediated by the maximum quit time across substances and differed by varying levels of DD.

**Conclusion:** Results suggest that longer quit time of *any* substance is related to improved recovery outcomes, particularly for individuals with low discounting rates. Together, interventions that focus on harm reduction and/or those that modulate DD may lead to improved clinical outcomes (including quit time and PrR) in individuals with a history of polysubstance use.

## 1. Introduction

In 2023, 7.5 million US residents met DSM-5 criteria (American Psychiatric Association, 2013) for two or more substance use disorders (SUDs) simultaneously (including alcohol and other drugs) in the past year (SAMHSA, 2023). Moreover, polysubstance use has been associated with nearly half of the overdose deaths and worst treatment outcomes (Crummy et al., 2020). These statistics highlight the prevalence (Agrawal et al., 2007) and growing trend towards polysubstance use due to the ability to produce enhanced euphoric effects and overall pleasurable experiences (Boileau-Falardeau et al., 2022). Although SUDs are typically categorized by identifying the primary substance used, and multiple-SUDs or poly substance use disorder (polySUD) is not in the DSM-5 manual (Hasin et al., 2013), there is growing evidence of increased prevalence of polySUD in nationally representative samples, clinical samples, and in samples of individuals seeking treatment for problematic substance use (Brooner et al., 1997; Erga et al., 2021; Palamar et al., 2018; Staines et al., 2001; Timko et al., 2018). Here, we define polySUD as the individual meeting DSM-5 criteria for two or more SUDs during their lifetime.

Evidence suggests that polysubstance use is increasingly prevalent (Agrawal et al., 2007; Eddie et al., 2022; Grant et al., 2016; Mefodeva et al., 2022). For example, individuals who use alcohol frequently also use other drugs, such as cocaine, cannabis, benzodiazepines, and heroin (McCabe et al., 2006; Moss et al., 2015; Petry, 2001). Further, polysubstance use is associated with worse SUD treatment access and outcomes (Bhondoekhan et al., 2023; Frost et al., 2021; Krawczyk et al., 2021; Mackay et al., 2021). Despite the high prevalence and potentially severe consequences from polysubstance use, mono-drug use has been more extensively investigated (Bamsey, 2017; McCabe et al., 2006; Mefodeva et al., 2022; Schensul et al., 2005).

Additionally, polysubstance use is often an exclusion criterion in clinical studies (Amlung et al., 2017). While there have been studies focused on polysubstance use, they have been few and far between with variable criteria for polysubstance use (Bailey and McHugh, 2023; Bhalla et al., 2017; Bhondoekhan et al., 2023; Eddie et al., 2022; Goodwin et al., 2022; Grant et al., 2016; McCabe et al., 2017). Hence, a noticeable gap exists in recovery and epidemiological research regarding polysubstance use behavior (McCabe et al., 2006).

An important aspect when considering recovery success from polysubstance use is whether to focus on recovery from the primary substance or all substances (Brecht et al., 2008). If recovery success is to be measured in relation to all drugs, success rates may indeed be low for individuals with polySUDs (Erga et al., 2021). A better approach might be to look at recovery from polysubstance use not as an all-or-none phenomenon, but as allowing for degrees of variation. That way, we do not invalidate behavior change and progress in one substance by the lack of change from another. Additionally, myriad reasons exist why someone may seek treatment/recovery from some but not all addictive substances, including not identifying all addictive behaviors as problematic or having direct health impacts from one substance but not the other (Kelly et al., 2018). While these ideas have been acknowledged, the current approach to treatment and recovery is static, that is, focused on one primary substance. However, this perspective is disconnected from the current polysubstance use landscape, and our focus should now be redirected to approaches that encompass and reflect recovery progress. While the traditional all-or-none approaches are still valid, history and/or concurrent polysubstance use must be considered. Therefore, we propose the notion of proportion of remission (PrR) as an additional and potentially more sensitive measure of recovery success from polysubstance use. We define PrR as the proportion of substances an individual is in remission from in the past 12-months relative to the total number of lifetime SUDs, as measured by DSM-5 (i.e., number of substances in remission divided by the total number of lifetime substances).

Another measure that has been shown to be robustly related to addiction is delay discounting (DD) - an index of the extent to which an individual prefers immediate reinforcement over larger reinforcement later (Amlung et al., 2017; Bickel et al., 2021; MacKillop et al., 2011). DD is a well-established predictor and behavioral marker of SUD and other behavioral disorders, and has been shown to predict drug use, treatment outcomes, and remission (Athamneh et al., 2022c; Martínez-Loredo et al., 2018; Moody et al., 2016). DD rates are associated with substance use initiation (Audrain-McGovern et al., 2009), severity (Albein-Urios et al., 2012; Johnson et al., 2007; Vuchinich and Simpson, 1998), and time in recovery (Athamneh et al., 2019; Craft et al., 2021; Tomlinson et al., 2020). Indeed, one of the current theories of addiction, Reinforcer Pathology, states that addiction results from a short temporal window (i.e., high DD) which increases the valuation of brief, intense, and immediate reinforcers (e.g., drugs) to the detriment of reinforcers that accrue value over time (e.g., prosocial reinforcers) (Bickel et al., 2014a; Bickel and Athamneh, 2020). Importantly, previous studies have shown that DD rate is a strong predictor of recovery success, including remission and improved quality of life (Athamneh et al., 2023, 2022c).

Most studies investigating DD and SUD focus on a single or primary SUD, despite the high prevalence of polysubstance use (Bailey and McHugh, 2023). Previous research that considered multiple substances found that individuals that use two substances discount significantly more (i.e., higher DD) than those who use one substance (Moody et al., 2016). However, this study was limited to individuals that use one, two, or three substances, and the analysis restricted to alcohol, cocaine, and tobacco use. Thus, the relationship between DD and polysubstance use amongst individuals in recovery is relatively understudied. Further, to our knowledge, research on associations between DD and proportion of remission has yet to be explored. Furthermore, even less is known about the recovery trajectories for individuals with lifetime polysubstance use disorders.

Therefore, the primary objective of this study was to examine the association between PrR and key dimensions of SUD and recovery exclusively in individuals in recovery from polySUDs. Specifically, the goal was to examine associations between PrR and time in recovery, time since quitting each substance, and DD. We hypothesized that greater time in recovery, greater quit time, and lower DD would be associated with a higher PrR (i.e., greater number of substances one is in recovery from relative to the number of lifetime SUDs). Further in an exploratory analysis, we examined whether maximum quit time (M) mediated the relationship between time in recovery (X) and proportion of remission (Y), and whether each path was moderated by DD (ln(*k*); W). We used time in recovery (i.e., time since first attempt to change substance using behavior) as the X and maximum quit time (i.e., time since cessation of substance use) as the M because initiating behavior change naturally precedes ceasing substance use. Further, we decided to include DD as a potential moderator of each path (see **Supplementary Figure 1** for a conceptual model) given literature that shows a relationship between DD and time in recovery (Craft et al., 2021), intention to quit (Athamneh et al., 2017) as well as abstinence (MacKillop and Kahler, 2009; Sheffer et al., 2012, 2014) and SUD remission (Athamneh et al., 2023, 2022b, 2022c).

## 2. Methods

### 2.1 Participants

Participants (N = 2,417) were recruited from an online recovery registry, the International Quit and Recovery Registry (IQRR; www.quitandrecovery.org). For additional details regarding the IQRR, including the assessments, compensation structure, and recovery community, see (Athamneh et al., 2022c; Craft et al., 2021). To be part of IQRR, participants are required to complete a baseline assessment that collects demographic information, personal history of SUD, and multiple dimensions of recovery such as DD and time in recovery. All data was collected using two online survey platforms: QuestionPro (Austin, TX) and Qualtrics (Provo, UT). The data used in this study were collected between September 2019 and June 2023. This study was approved by the Institutional Review Board at Virginia Tech. Inclusion criteria for this secondary analysis were: 1) 18 years or older, 2) met lifetime DSM-5 criteria for two or more substances, and 3) self-reported to be in recovery from at least one of their lifetime SUDs.

### 2.2 Measures

#### 2.2.1 Demographics

Demographics, including personal (i.e., age, gender, ethnicity, race) and socio-economic variables (i.e., education, combined household income) were collected.

#### 2.2.2 Lifetime substance use

Participants were asked about substance use in their lifetime that was either to get high or self-medicate a problem without the approval of a doctor. The substances were 11 broad categories, including nicotine, alcohol, cannabis, opioids, cocaine, stimulants, prescription pain relievers, hallucinogens, tranquilizers, depressants, and inhalants. Consistent with DSM-5, individuals who endorsed at least 2 symptoms of a substance were considered to have met lifetime history for that SUD.

#### 2.2.3 Recovery

For each substance that participants met lifetime criteria for, a follow-up question regarding whether they have ever tried to recover/initiate recovery from that substance was asked. Here, we take a naturalistic perspective on recovery; initiating recovery means that they started actively changing their substance use behavior and doing things to cut down or stop using (not just thinking about changing their substance use) whether or not they were successful.

#### 2.2.4 SUD Remission status

Remission status was assessed for each SUD an individual met lifetime criteria for. Individuals who endorsed 1 or more DSM-5 criteria (excluding craving, per DSM-5) in the past 12 months were considered *not in remission* (Hasin et al., 2013). Conversely, individuals who did not endorse any symptom of that SUD in the past 12 months, excluding craving, were categorized as *in remission*.

#### 2.2.5 Proportion of remission (PrR)

PrR represents the number of substances in 12-month remission divided by the total number of lifetime SUDs. Here, this is encoded as a binomial variable indicating the number of substances an individual is in remission (i.e., successes) and the number of substances an individual is not in remission (i.e., failures). Note that the total number of substances considered varied across participants based on their lifetime history.

#### 2.2.6 Maximum time in recovery

Participants reported the date they first initiated an attempt to change their substance using behavior - termed their recovery date - for each SUD. For each SUD, their time in recovery was calculated as the number of days between their recovery initiation date and the date they completed the survey. Their maximum time in recovery was considered the longest time since initiating recovery for any reported substance. For example, an individual with a history of two lifetime SUDs (e.g., alcohol and cocaine) who initiated recovery from alcohol 2.5 years ago and from cocaine 1.35 years ago was considered to have a maximum time in recovery of 2.5 years.

#### 2.2.7 Maximum quit time

Participants reported the date of last use for each substance they met lifetime DSM-5 SUD for (i.e., a substance-specific quit date). *Quit time* was calculated for each substance by taking the number of days between their reported date of last use of that substance and the date they completed the survey. Participants could have different quit times for each lifetime SUD. The maximum quit time was considered for the substance with the longest time since last use.

#### 2.2.8 Delay discounting

Participants completed a $1000 adjusting amount DD task (Du et al., 2002). Specifically, participants made choices between a smaller sooner (SS) and a larger later (LL) monetary rewards for six time points (1 day, 1 month, 6 months, 1 year, 5 years, 25 years) presented in a random order (Athamneh et al., 2022b, 2022c, 2019; Craft et al., 2021). Participants completed six trials for each time point. The amount of the SS was titrated up or down depending on the participant’s choice in the previous trial. Responses for each time point were used to calculate the indifference point (i.e., the point at which the value of the SS rewards is subjectively equivalent to that of the LL). Discounting rate (k, log-transformed for analysis to normalize the data and stabilize variance) was calculated using Mazur’s hyperbolic equation (Mazur, 1987).

### 2.3 Data cleaning

Completed baseline responses (n=2,417) collected between September 2019 and June 2023 were included in the final sample. In addition to our inclusion criteria detailed above, we also excluded participants who: 1) indicated a date of last use (i.e., quit date) or recovery initiation date more than two days in the future (we considered a two day window to allow for a margin of error); 2) indicated a longer maximum quit time than their maximum time in recovery (i.e., potentially unsystematic responders); 3) provided non-systematic delay discounting data (Johnson and Bickel, 2008).

### 2.4 Statistical Analysis

All statistical analyses were conducted using R software (version 4.2.2) (R Core Team, 2022). The analysis was not pre-registered and should be considered exploratory. Participant characteristics were described using mean (standard deviation) and frequencies, where appropriate. The proportion of remission was modeled using binomial regression at a group level; that is each substance for which a participant met lifetime criteria was a Bernoulli trial with remission equaling 1 and not in remission equaling 0. The total number of substances (i.e., Bernoulli trials) considered varied across participants based on their lifetime history. The binomial regression was modeled at a group level. We performed multivariate binomial regressions to assess the association of proportion of remission with our variables of interest (i.e., delay discounting, maximum time in recovery, and maximum quit time) and demographics. Maximum time in recovery and maximum quit time were log-transformed to assure normality. Results from multivariate binomial regressions were reported as adjusted odds ratios and 95% confidence intervals (CI). Additionally, effect sizes were calculated for all variables and reported as Cohen’s *f*. Significance level was defined as p < .05 for all analyses.

To further understand the relationship between proportion of remission from polysubstance use and our set of recovery measures, we conducted a moderated mediation. We performed bootstrapping to estimate 95% confidence intervals (CI) using 10,000 bootstrap samples. A significant indirect effect is established if the 95% CI of the coefficient for the indirect path does not include zero (Preacher et al., 2007). Moderation was further probed by estimating and plotting the indirect effects of time in recovery at values of discounting corresponding to the 16th, 50th and 84th percentile points. These three points represent low (W = -7.787), moderate (W = -5.409) and high (W = -2.681) values of discounting in the current sample.

## 3. Results

### 3.1 Participant demographics and recovery characteristics

Participant characteristics of the total analytical sample (n=2,417) are described in **Table 1**. Mean age of participants was 41.06 years old, with 58.6% female, 77.6% White, and 91.5% non-Hispanic. The median maximum time in recovery was 7.14 years while median maximum quit time was 5.28 years. Notably, 17.17% of the sample had two, 16.79% had three, and 66.04% had four or more lifetime substances based on DSM-5 criteria. The most common substances of use were alcohol (85.14%), tobacco (80.17%), and cannabis (67.8%).

**Table 1:** Participant characteristics.

|  | Overall |
| --- | --- |
| n | 2417 |
| Age (mean (SD)) | 41.06 (11.93) |
| Education (mean (SD)) | 14.21 (3.96) |
| Gender = Male (%) | 1000 (41.4) |
| Race (%) |  |
| American Indian or Alaska Native | 40 (1.7) |
| Asian | 17 (0.7) |
| Black or African American | 352 (14.6) |
| Multiracial | 78 (3.2) |
| Native Hawaiian or Other Pacific Islander | 5 (0.2) |
| Other | 50 (2.1) |
| White/Caucasian | 1875 (77.6) |
| Ethnicity = Not Hispanic or Latino (%) | 2211 (91.5) |
| Household income (%) |  |
| <19.99k | 704 (29.1) |
| 20k-34.99k | 351 (14.5) |
| 35k-69.99k | 576 (23.8) |
| >70k | 786 (32.5) |
| Marital status (%) |  |
| Married/Cohabiting | 1048 (43.4) |
| Not Married | 866 (35.8) |
| Separated/Widowed/Divorced | 503 (20.8) |
| Region (%) |  |
| Midwest | 472 (19.6) |
| Northeast | 418 (17.4) |
| South | 980 (40.7) |
| West | 538 (22.3) |

### 3.2 Predictors of proportion of remission

We performed multivariate binomial regression predicting PrR as a function of DD, maximum time in recovery, and maximum quit time, each adjusted for demographic variables (Figure 1). Relative to individuals with high DD rates (shorter temporal window), those with high DD showed 14% reduced odds of achieving a higher PrR (i.e., being in remission from more substances relative to lifetime SUDs). Specifically, each unit increase in DD was associated with 14% lower odds of higher PrR (aOR: 0.86; 95% CI: 0.85, 0.88; p < .001). Additionally, individuals with greater maximum time in recovery across lifetime substances had a greater likelihood of being in a higher PrR [aOR: 1.75; 95% CI: 1.67, 1.84; p <.001). Similarly, individuals with greater maximum quit time across substances were twice as likely to be in a higher PrR [aOR: 2.1, 95% CI: 2.01, 2.20; p <.001). See **Table 2** for complete model results.

**Figure 1:**
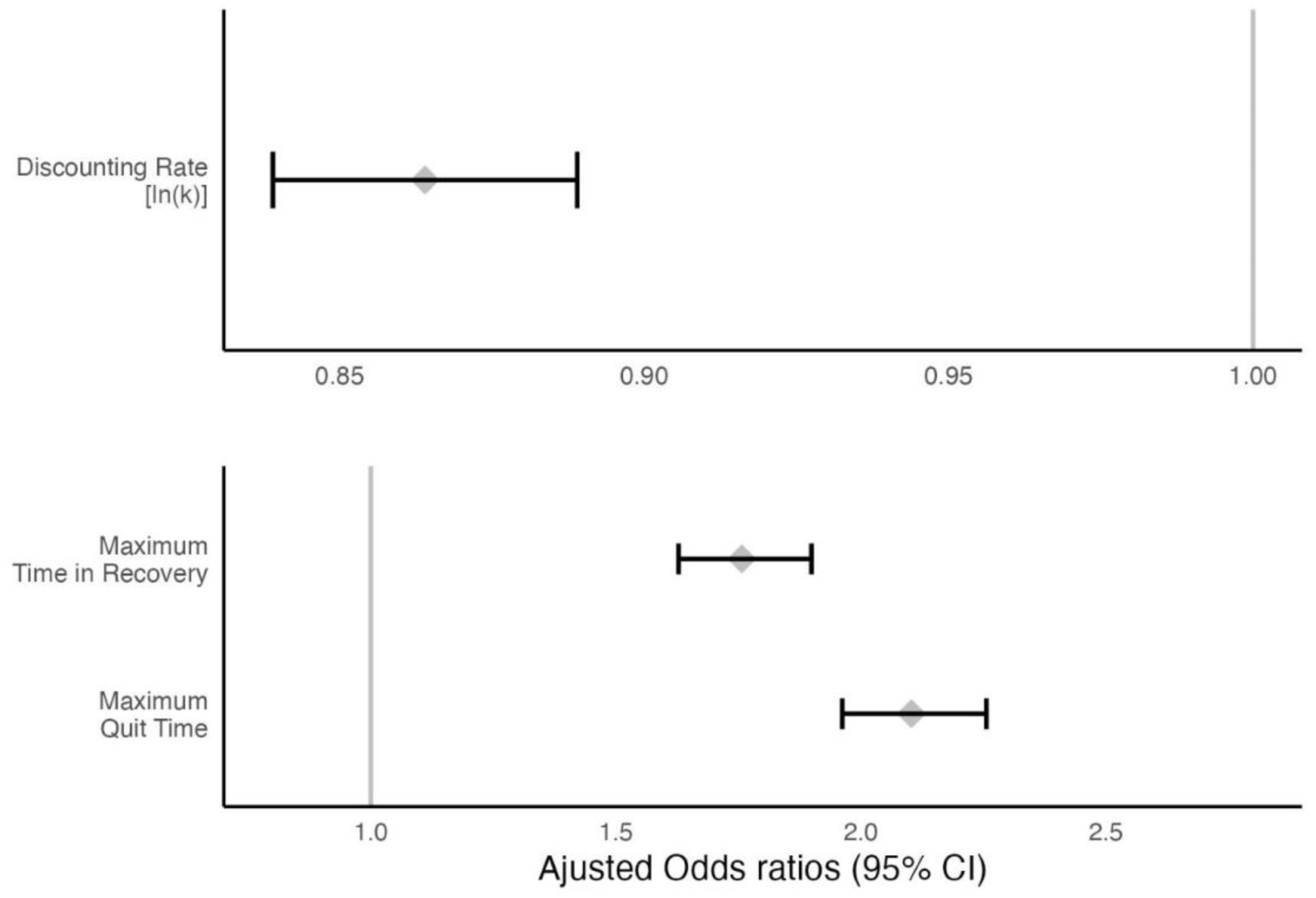
Multivariate binomial regression examining the association between delay discounting (DD), maximum time in recovery, maximum quit time, and the proportion of remission (PrR), adjusted for demographic variables.

**Table 2.**
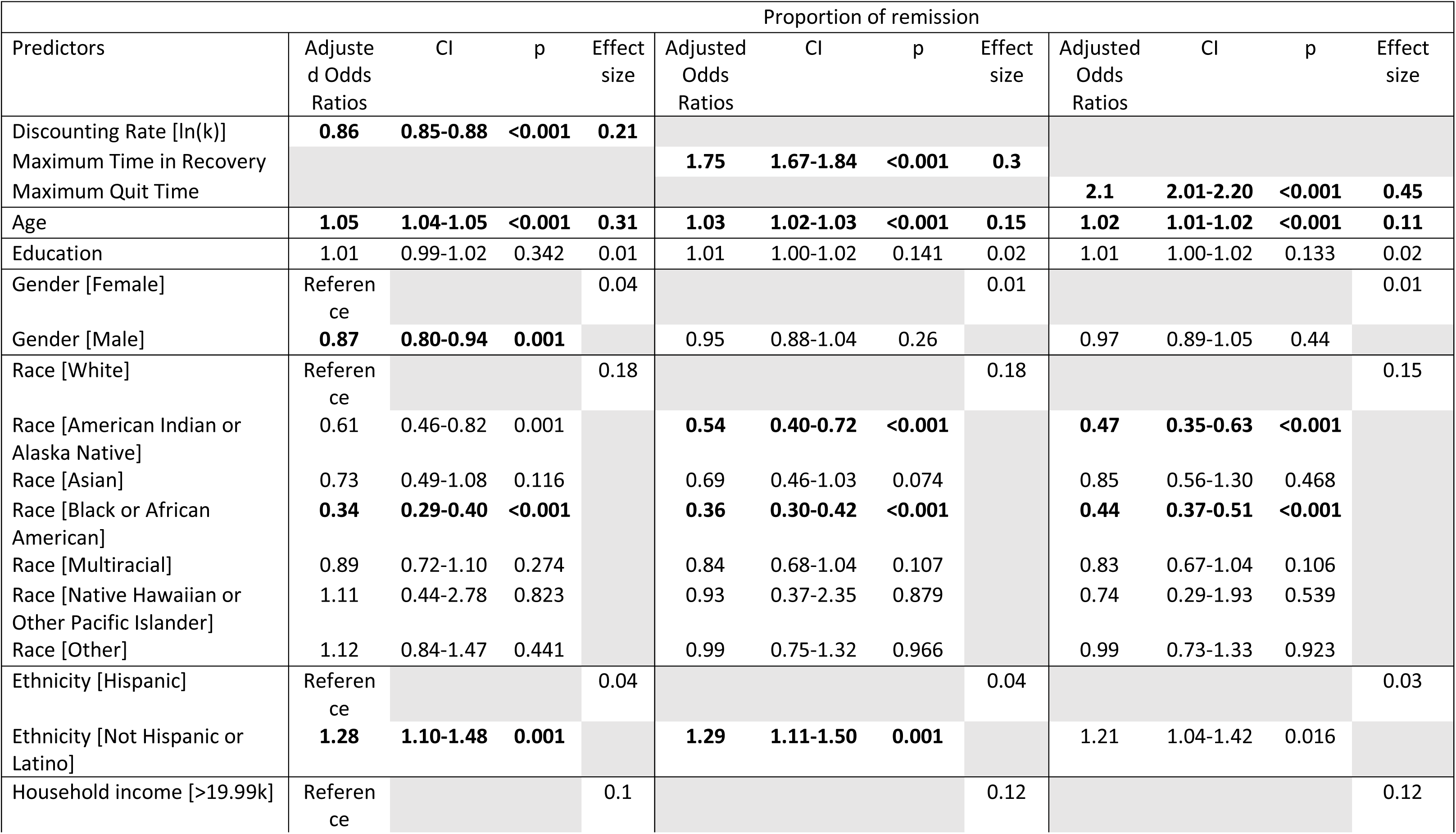
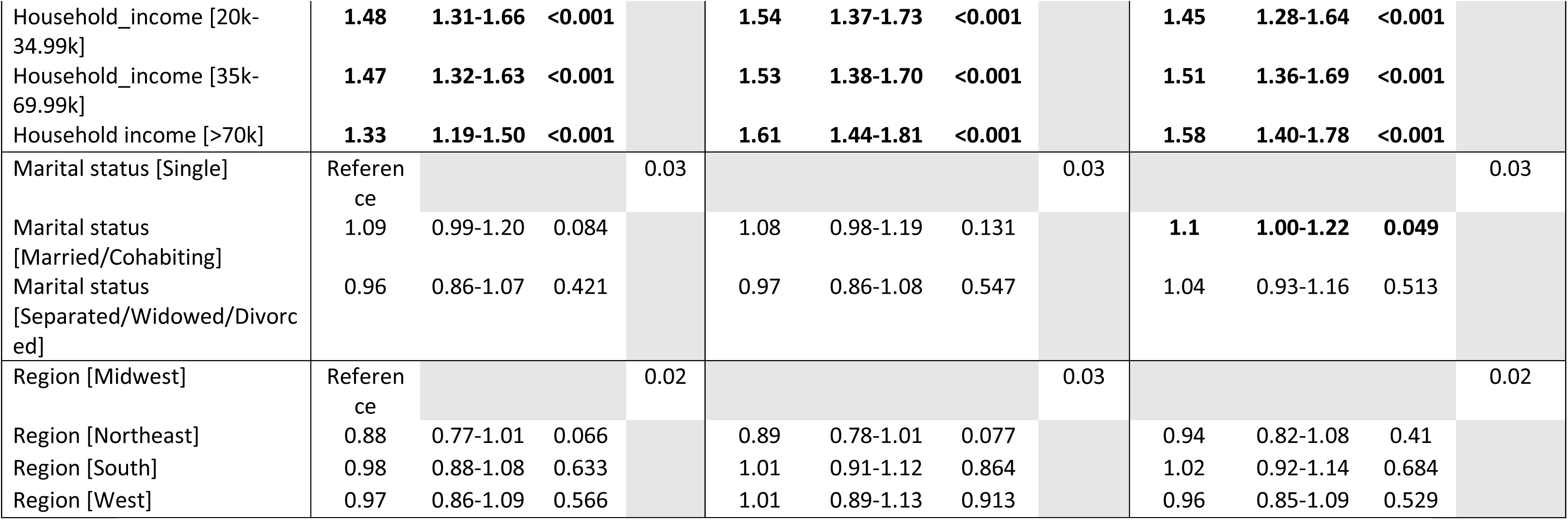
Multivariate binomial regression for variables of interest along with demographics.

### 3.3 Moderated Mediation Analysis

A moderated mediation analysis was performed to further understand the relationship between DD rate, maximum time in recovery, and maximum quit time, and how these variables impact PrR. First, a significant indirect effect between maximum time in recovery and proportion of remission (PrR) was mediated by the maximum quit time across substances (coefficient = 0.739, 95% CI = 0.459, 1.040; Figure 2A). Second, this mediation was moderated by delay discounting. Specifically, individuals with higher discounting (i.e., more impulsive choices) had weaker associations between time in recovery and quit time (W → M: coefficient = –0.150, 95% CI = –0.233, –0.070), with further moderation via the interaction of time in recovery and delay discounting (XW → M: coefficient = –0.016, 95% CI = –0.028, –0.004). Third, the direct effect of time in recovery on PrR was non-significant (coefficient = 0.091, 95% CI = -0.212, 0.378), suggesting the indirect effect explains a relationship between time in recovery and PrR. Figure 2B illustrates the indirect effect at three observed percentiles of delay discounting (ln(k)) — at the 16th (W = –7.787), 50th (W = –5.409), and 84th (W = –2.681) — with confidence intervals that do not include zero, confirming the significance of the moderated indirect effect across these levels.

**Figure 2:**
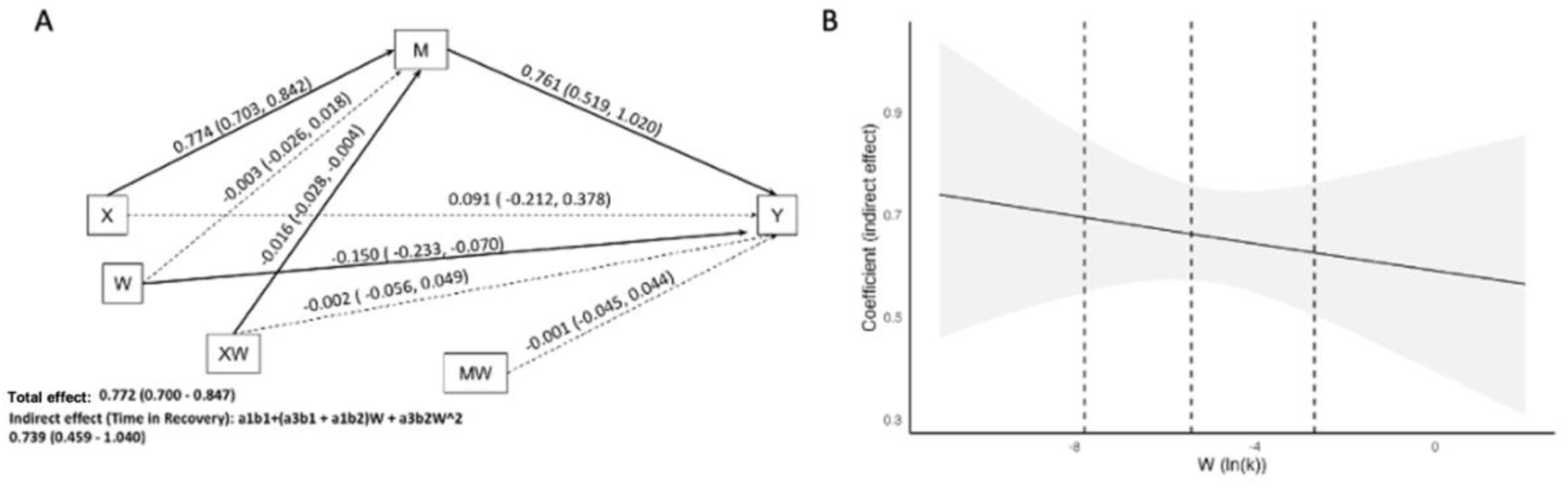
A) Moderated mediation analysis testing whether maximum quit time (M) mediates the relationship between time in recovery (X) and proportion of remission (Y), with delay discounting (ln(k); W) as a moderator of each path in the model. B) Indirect effects of maximum time in recovery on proportion of remission at three levels of delay discounting (ln(k)): 16th percentile (W = −7.787), 50th percentile (W = −5.409), and 84th percentile (W = −2.681). Confidence intervals for all estimates exclude zero, indicating statistically significant mediation.

## 4. Discussion

The present study examined the proportion of remission (i.e., the number of substances an individual is in 12-month remission from divided by the number of lifetime SUDs for which they met DSM-5 criteria) amongst a sample of individuals in recovery from addiction. We found that longer reported maximum time in recovery and maximum quit time (i.e., longest abstinence interval from any given substance) across substances was associated with a higher proportion of remission. In addition, we report for the first time that individuals with a high discounting rate (i.e., shorter temporal window) with a lifetime history of polySUD had 14% lower likelihood to be in higher proportion of remission than individuals with lower discounting rates (i.e., longer temporal window). We also found that the maximum quit time mediated the relationship between time in recovery and proportion of remission, with delay discounting moderating the relationship. Hence, our findings add to the growing evidence of discounting as a marker of substance use pathology.

We found that longer time in recovery and maximum quit time was associated with a higher proportion of remission. Indeed, prior research has demonstrated that quitting one substance has been related to improved outcomes for other SUDs (Nguyen et al., 2020). Nguyen et al., (2020) found that in a sample of Veterans with alcohol use disorder (AUD), individuals who smoked previously were 6.6 times more likely to achieve 6-month abstinence from alcohol compared to those who smoked currently, and individuals with past smoking were also more likely to achieve 6-month alcohol abstinence compared to individuals who never smoke. This study also found that more abstinent days pre-treatment were also related to improved treatment outcomes (Nguyen et al., 2020). Similarly, improved QoL and executive function, as well as lower psychological distress, among abstinent individuals compared to individuals who had returned to use has also been reported (Hagen et al., 2017). A possible explanation for this finding is that individuals who have been able to abstain from one substance for longer may have better self-regulatory skills, fewer physical health problems, less psychiatric comorbidity, and improved neurocognitive functioning, potentially contributing to a higher proportion of remission.

We also found that delay discounting was significantly associated with PrR and moderated the indirect relationship between maximum quit time and PrR. That is, lower delay discounters with higher maximum quit time exhibited a higher proportion of remission compared to higher delay discounters. This finding aligns with prior research demonstrating higher levels of delay discounting in individuals with polysubstance use compared to those with single substance use (tobacco only; Moody et al., 2016; Naudé et al., 2022). Our results are also consistent with a robust body of research showing that lower delay discounting is associated with remission (Athamneh et al., 2023, 2022b, 2022c; Bickel et al., 1999; Craft et al., 2022), lower rates of relapse following an abstinence period (González-Roz et al., 2019), better treatment outcomes (Loree et al., 2015), and recovery progress (Craft et al., 2021). Conversely, higher delay discounting is associated with worse SUD severity (Albein-Urios et al., 2012; Amlung et al., 2017; Johnson et al., 2007; MacKillop et al., 2011). Craft et al., (2021) found that 1) delay discounting mediated the relationship between time in recovery and recovery progress and 2) a majority of individuals in self-reported recovery considered abstinence important for successful recovery. Together, these findings provide further support that measurable reductions in any substance use (i.e., quitting any of multiple substances of abuse) and improvements in underlying etiological factors of SUD (i.e., delay discounting) may better capture positive behavior change and successful recovery compared to complete abstinence, complete remission, or time in recovery.

Clinically, interventions that modulate delay discounting may be particularly effective in lengthening quit time and thus increasing proportion of remission in individuals with polysubstance use. For example, episodic future thinking (EFT) has been shown to reduce delay discounting in individuals with AUD (Athamneh et al., 2022a; Craft et al., 2023; Voss et al., 2022), cocaine use disorder (Snider et al., 2021), individuals who smoke (Chiou and Wu, 2017; Stein et al., 2016) as well reduce consumption across several substances (Athamneh et al., 2022a; Chiou and Wu, 2017; Stein et al., 2016). Our findings on the moderating effects of discounting are more broadly congruent with studies on rate dependence, a phenomenon in which post-treatment change is dependent on pre-treatment response level. Craft et al., (2023) found that individuals with higher discounting pre-EFT exhibited greater change in their discounting rates post-EFT compared to those with low discounting pre-EFT. Further, in a re-examination of other interventions (e.g., working memory training and contingency management), rate dependent changes in discounting were found and, the treatments with the largest effects on discounting also exhibited the most improvements in abstinence(Bickel et al., 2014b). Together, amongst individuals with a history of polysubstance use, those with high discounting rates and less maximum quit time may particularly benefit from interventions that modulate delay discounting.

This paper has several limitations that are important to acknowledge. First, this study was cross-sectional and therefore, it limits the interpretations to non-causal inferences (Maxwell et al., 2011). While our model is strengthened because time in recovery (i.e., years since initiating behavior change; X) naturally precedes longest abstinence interval (i.e., longest time since actually quitting use of a substance; M), future longitudinal research on the associations between maximum quit time, delay discounting, and proportion of remission would contribute to a better understanding of recovery in polysubstance use. Second, the generalizability of the results may be limited because the IQRR is an online registry and in this, individuals self-select to participate and must have access to the internet. Third, our conceptualization of polysubstance use considered the lifetime history of multiple SUDs and did not examine concurrent polysubstance use. Concurrent use of substances has been shown to be related to poor treatment outcomes (Lin et al., 2021; Moody et al., 2016; Schauer et al., 2017; Staiger et al., 2013) and future research on concurrent polysubstance use as it relates to recovery is warranted.

Despite these limitations, this paper also has several strengths that should be recognized. We conducted this research on a large, diverse sample. Further, as Amlung et al (2017) noted in their meta-analysis (Amlung et al., 2017), most single SUD studies had no exclusion criteria for other SUDs, thus likely representing polySUD cohorts. Our study included individuals who met lifetime SUD criteria for all 11 major substance categories represented in the DSM-5, and reported these SUD histories. Finally, our analysis was not limited to one pathway of recovery (e.g., sobriety) and rather, examined multiple aspects of SUD recovery - including self-reported behavior change, remission, and quit duration.

In this paper, we propose a novel construct of SUD recovery: the proportion of remission (i.e., the number of SUDs an individual is in remission from relative to the total number of lifetime SUDs). Our findings suggest that longer quit time of *any* substance for which an individual has a lifetime SUD history is related to improved recovery outcomes, particularly for individuals with low discounting rates. This suggests that positive behavior change may occur in individuals that use multiple substances who are able to reduce use of even one of their substances of abuse. To help support this perspective clinically, interventions that focus on harm reduction and/or those that modulate delay discounting may lead to improved clinical outcomes (including quit time and proportion of remission) in individuals with a history of polysubstance use.

## Data availability

Data will be made available on reasonable request.

## CRediT statement

FQ: Conceptualization, Formal analysis, Methodology, Writing – original draft

DCT: Methodology, Writing – review and editing

RMF: Writing – original draft, Writing – review and editing

ANT: Conceptualization, Project administration, Writing – review and editing, Supervision

WKB: Conceptualization, Project administration, Writing – review and editing, Supervision

