## Supplementary figures and images for "Polysubstance use: Delay discounting in relationship to remission"

### Supplemental

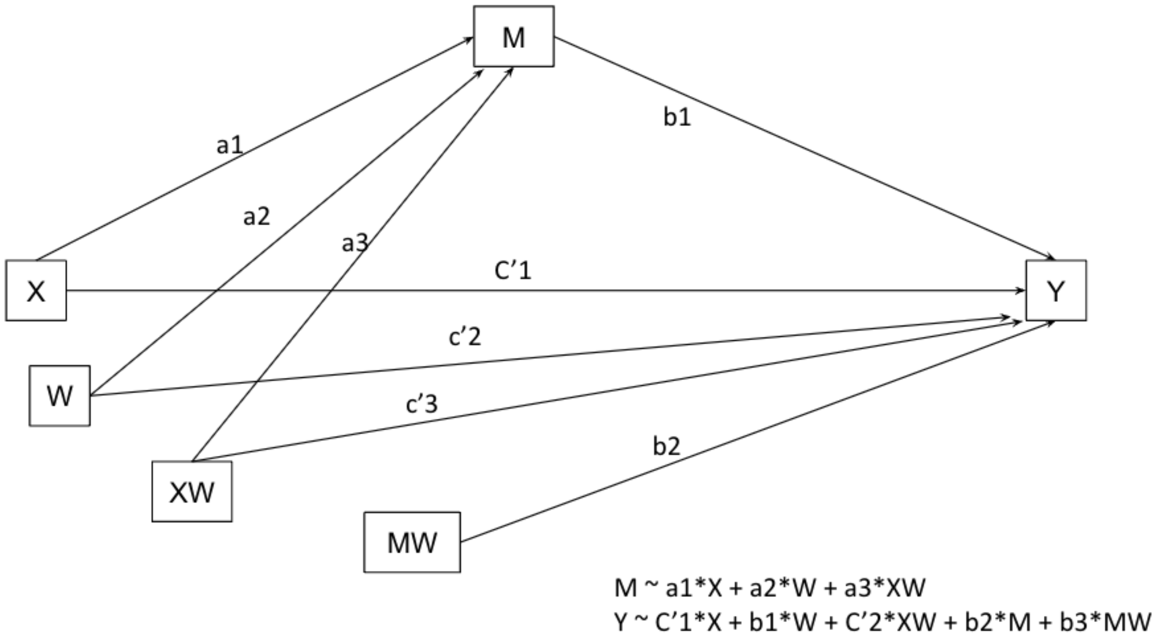


Supplementary Figure 1. Conceptual Moderated Mediation Model.
